# BREATH-PATH – Protocol for lung cancer management in surgical patients through analysis of breath profiles

**DOI:** 10.1101/2025.01.19.25320800

**Authors:** Rita Barata, Catarina Carvalheiro, Bernardo S. Raimundo, Marcos Pantarotto, Pedro D. Vaz

## Abstract

**Background:** Anatomical surgical resection is the treatment of choice for early-stage non–small cell lung cancer (NSCLC) and is a part of the multimodality treatment for resectable, locally advanced NSCLC. Local, regional and distant recurrence is the most common cause of treatment failure after resection. Multiple studies have recommended chest computed tomography (CT) for lung cancer (LC) patient’s follow-up. Even though adopting this strategy, the recurrence will only be detected months after the onset of the disease once imaging changes will only occur with a high tumor burden.

**Methods:** Breath analysis is a promising technology and useful addition to the currently available modalities to achieve lung cancer diagnosis and management. The volatile organic compounds (VOCs) produced as end-products of metabolism by the LC cells (and present on the exhaled breath of the patients) seem to have a unique pattern that can be used as a tool to detect lung cancer (VOC profile).

**Discussion:** The BREATH-PATH study aims to establish a VOC profile for all LC patients at diagnosis and understand its variations with the different implemented treatments in order to find response patterns. By understanding these patterns and identifying their deviations thereof with disease relapse supported by imaging methods we will be able to validate the breath analysis as a useful tool for LC recurrence detection.

This study intents not only to reiterate the methods’ efficacy but also to understand how high its sensitivity is in detecting a tumor recurrence, allowing breath analysis to be integrated into clinical decision algorithms.

## Introduction

Lung cancer is the leading cause of cancer morbidity and mortality worldwide, with almost 2.5 million new cases and over 1.8 million deaths in 2022. It is responsible for 12.4% of the cancers diagnosed globally and approximately one-fifth (18.7%) of cancer deaths. It also ranks first among men and second among women for both incidence and mortality, with a male-to-female lung cancer incidence and mortality ratio of around two(1).

Surgical resection is the primary treatment for early-stage presentations of non-small cell lung cancer (NSCLC), with recent evidence supporting sub-lobar resections (segmentectomies and wedge resection) as an option for patients with nodules smaller than 2 cm (IA1/IA2 - T1abN0). Surgical treatment is also considered for selected patients’ resectable, locally advanced (NSCLC- LA) stages(2–5).

Regardless of numerous advances in diagnosis, pathologic and molecular classification, and treatment options in the last few years, disease recurrence after treatment with curative intent remains a significant problem(6–13).

Patients with completely resected stage I-II NSCLC have a 5-year cumulative incidence of recurrence of 20.1%; fifty percent of these patients experience recurrences at distant sites and 31.9% at both locoregional and distant sites, with a median time to recurrence of 18.8 months(14). A considerable number of patients with stage I-III lung cancer will have local recurrence (22-50%) or distant recurrence (3-20%) following curative intent treatment(15).

Although the risks of recurrence and new primary lung cancer are well known, an optimal follow-up strategy has yet to be defined. A thoracic computed tomography (CT) scan is currently the recommended method of imaging for the follow-up of resected lung cancer patients according to European and North American guidelines(4,16). Although there is no evidence of an added overall survival benefit, serial CT scans during follow-up of resected NSCLC patients allow for earlier detection of recurrence, as well as new primary lung cancers(17–19).

A CT-scan only evaluates the solid and topographic cancer structure and morphology, but the actual concept of tumor is much more comprehensive, being defined as a disease that has its own genetic, biological and metabolic identity(20). It is based on this new assumption that the exhaled breath analysis arises as a promising noninvasive method for detecting and managing lung cancer. Exhaled breath contains a complex mixture of non-volatile and volatile organic compounds (VOC) produced as end-products of cellular metabolism. VOCs are excreted by diffusion and exhaled through breath. All pathophysiological variations at the intracellular level have consequences for cellular metabolism and thus on the excreted products, including VOCs. Therefore, changes in the VOC profile are expected throughout the disease’s natural history, in either response to treatment or disease progression(20–28).

Developing noninvasive tests accessible at the point of care could significantly change clinical decision-making improving disease outcome. Because breath tests often have high negative predictive values (NPV), a positive VOC test may aid doctors in developing a more straightforward and less expensive pathway for earlier diagnosis of new or recurrent tumors(24).

With this protocol, we aim to demonstrate 1) a positive VOC pattern for patients with local and LA-NSCLC; and 2) changes in VOC patterns throughout the surgical treatment of patients with local and LA-NSCLC, establishing the foundations for using VOC as a tool in the future follow-up of patients with resected lung cancer(22).

## Material and methods

BREATH-PATH is a prospective, single-center, observational study aiming to identify changes in the VOC profile of patients recently diagnosed with stages I-IIIB NSCLC (TNM 9th edition) who are undergoing surgical treatment, whether alone or in combination with neoadjuvant, adjuvant, or peri-operative therapies throughout the postoperative follow-up. Its objective is to determine whether the VOC profile can serve as a tool for predicting lung cancer recurrence in the future. This study builds upon the recent findings of a previous study carried out at the same institution by the same team(26). The study showed that analyzing VOC profiles from exhaled breath is a practical and feasible method for high-accuracy LC detection.

The study was reviewed and approved by the Champalimaud Foundation Institutional Review Board.

Recruitment commenced on 16 July 2024, as per the ethics committee approval letter. A population of 50 adult patients is anticipated to be recruited from the lung cancer unit at the Champalimaud Foundation in Lisbon, Portugal, for approximately 15 months. The study will conclude after *circa* 40 months of the last patient enrollment.

The patients included in the study will follow international guidelines for diagnosing, staging, treating, and following up on their lung cancer. All clinical decisions throughout the patient′s journey will be made in a multidisciplinary meeting.

Verbal and written information about the study will be provided to all participants and their inclusion in the study will only occur after their written consent is obtained.

### Inclusion criteria

1. Adult patients aged between 18 years or older and capable of signing the study’s informed consent;
2. Treatment naïve patients with a diagnosis of NSCLC at clinical stage IA – IIIB (according to TNM 9^th^ edition), considered resectable after multidisciplinary team meeting evaluation;
3. Patients must have respiratory function tests compatible with anatomical lung resection;
4. No previous diagnosis of neoplasia in the last five years, except for prostate cancer treated in the previous 3 years with curative intention, not dependent on hormonal treatment and without evidence of recurrence.

### Exclusion criteria

1. Any active clinical contraindication for surgery;
2. Inability to complete breath sampling procedure due to, e.g. hyper- or hypoventilation, diagnosis of neuromuscular disease, tracheostomy, respiratory failure, or claustrophobia when wearing the sampling mask;
3. Participation in a clinical trial of investigational medical products;
4. Active use of psychotropic drugs.

### Surgical requirements

Only anatomical surgeries will be accepted, such as segmentectomy, lobectomy, or pneumonectomy, in association with systematic mediastinal lymphadenectomy. According to the surgeon’s choice, the approach method may vary with video-assisted, robotic-assisted, or open thoracotomy. The completeness of the resection is defined by: (a) resection margins microscopically free of tumor; (b) systematic mediastinal lymphadenectomy, with at least three mediastinal stations resected, one of which is station 7; and (c) absence of extracapsular lymph node extension. Therefore, the completeness of resection is classified as R0 (no residual tumor), R1 (microscopic residual tumor), or R2 (macroscopic residual tumor).

### Groups Stratification

Allocation of patients to different groups depends on the treatment modality established according to the stage of the disease. Therefore, patients will be included in one of four groups:

1. **Group S** - Patients submitted only to surgery
2. **Group** _**N**_**S** - Patients submitted to neoadjuvant treatment (ChT, ChT + ICI) followed by surgery
3. **Group S**_**A**_ - Patients submitted to surgery followed by adjuvant treatment (ChT, ChT + ICI, Ch + TKI or TKI)
4. **Group** _**N**_**S**_**A**_ - Patients submitted to surgery and perioperative treatment (ChT + ICI / ICI)

All patients enrolled in this study will provide a first breath sample prior to any treatment—VOC N_BL_ (VOC before neoadjuvant treatment) or VOC S_BL_ (VOC before surgical treatment), depending on the group to which they are assigned, establishing a baseline VOC profile. Subsequent breath analyses will be done one month after surgery (VOC _P_S), three months after surgery (VOC_3_), and then every three months during the following two years (VOC_6_ to VOC_24_) or until disease recurrence, whichever occurs first. Patients will be stratified as shown in Fig 1.

**Fig 1.**
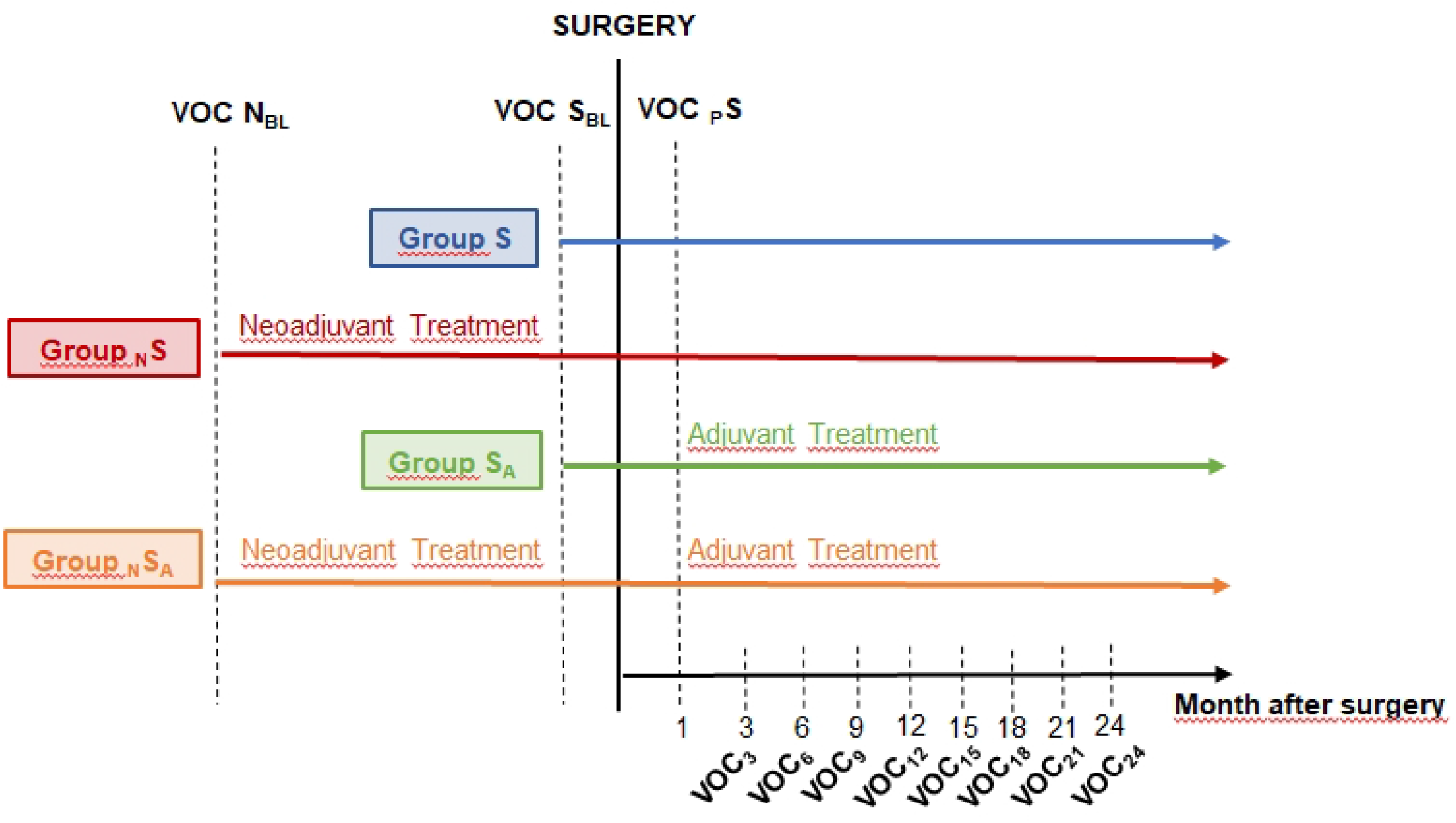
Study protocol: **Group S** - group of patients submitted only to surgery; **Group** _**N**_**S** - group of patients submitted to neoadjuvant treatment followed by surgery; **Group S**_**A**_ - group of patients submitted to surgery followed by adjuvant treatment; **Group** _**N**_**S**_**A**_ - group of patients submitted to surgery and perioperative treatment; **VOC N**_**BL**_ - VOC neoadjuvant baseline, before the beginning of any treatment; **VOC S**_**BL**_ - VOC surgery baseline, before surgery; **VOC** _**P**_**S** - VOC one month after surgery; VOC_3-24_ - VOC every three months after surgery during two years.

### Breath sampling collection and analysis by Gas chromatography- Field-asymmetric ion mobility spectrometry (GC-FAIMS)

Breath samples will be collected at Champalimaud Clinical Center.

To collect exhaled breath air, all enrolled volunteer patients must fast (food and any drinks apart from water) for at least four hours before sampling to avoid possible effects of food or its metabolites on the profile of volatile compounds. Volunteers cannot smoke or have oral hygiene for at least two hours before breath sampling, while prescription medicines should not be taken up to six hours before the procedure test. Collection of breath samples will be conducted directly with a mask holding thermal desorption (TD) tubes (Biomonitoring, inert coated tubes, Markes International Ltd). Clean air will be continuously supplied to the volunteer’s mask through a pump connected to an activated charcoal scrubber (CASPER system; Owlstone Medical). Subsequently, the ReCIVA breath sample system enables a completely non-invasive breath sampling process, allowing volunteers to perform normal tidal breathing. Through the software control, the ReCIVA system collects 2 L of full tidal exhaled breath, which is transferred onto the TD tubes at a flow rate of 500 mL/min.

All samples will be analyzed using a Gas Chromatography – Field Asymmetric Ion Mobility Spectrometry (GC-FAIMS) Lonestar instrument (Owlstone Medical, Cambridge, UK). FAIMS separates different ions in the gas phase based on their mobility across an oscillating electrical field related to shape and mass. This study will use the following settings: the dispersion field (DF) will be varied between 0 and 100% in 51 steps, and the compensation voltage (CV, to regulate which ions) will range from − V to +6 V.

The analysis of exhaled breath air using GC-FAIMS yields a complex yet information-rich dataset arising from the VOC analysis of the samples. This translates into a plot characterized by a pattern resembling a “chemical fingerprint” (the chemical composition of VOCs) for a given sample. The overall chromatographic profiles, devoid of qualitative information, will be classified by an AI algorithm previously established by the team and described in detail elsewhere(26).

### Patient consent

All eligible patients will receive an informed consent in plain language explaining the study’s key points, purpose, risks, and benefits. The doctors responsible for the study will address any questions patients may have concerning their participation.

The doctors and the patient will date and sign the consent form. Data will be collected using pseudo-anonymization, and patients will be assigned numbers according to their order of inclusion in the study.

### Data collection and handling

Data collected and its treatment will be accessed only by the team members involved in the study. Data to be collected will include the following: age, gender, smoking history, comorbidities, family history of neoplasia, lung cancer clinical and pathological stage, imaging and metabolic scan results, diagnosis and staging methods results, neoadjuvant and adjuvant treatments and their complications, surgical procedures and related data (morbidities, drainage time, length of hospital stay), and follow-up imaging test results. Data will be collected from the clinical processes, exams, and interventions of the patients during their clinical evaluation pathway.

The PI will be responsible for the pseudonymization of patients and for curating the data collected at the time of enrollment and throughout the duration of the study. All data collected will be handled exclusively at the Champalimaud Foundation.

### Planned statistical analysis

Statistical analyses of background data will be performed using descriptive statistical methods. For analytical inference, an alpha of 0.05 will be applied. All analyses will be conducted using SPSS software (IBM Corp. Released 2023. IBM SPSS Statistics for Windows, Version 29.0.2.0 Armonk, NY: IBM Corp).

Initial data processing of the exhaled breath chemical analysis will follow standardized procedures for handling complex data types, including corrections for experimental variability (baseline and time alignment). After pre-processing, chromatograms will be integrated to obtain the respective Area Under the Curve (AUC) and normalized to the unit area by division by the respective AUC.

Classification of the breath VOC profiles will involve filtering raw data for irrelevant features. This procedure allows not only the removal of statistically irrelevant data but also a drastic reduction in the size of the dataset. Subsequently, the classification of classes will be conducted using a supervised multiparametric classification AI algorithm, as described previously (21). This analysis will yield the corresponding confusion matrix and receiver operating characteristic (ROC) plots, allowing for the evaluation of sensitivity, specificity, true positive and true negative rates, and accuracy of the method.

## Discussion

In the last decade, many advances have been made regarding treating lung carcinoma (LC) patients. Immune checkpoint inhibitors (ICIs) and tyrosine kinase inhibitors (TKIs), alone or in combination with chemotherapy (ChT), have brought significant improvements to the prognosis of lung cancer patients, with gains in event-free survival (EFS), disease-free survival (DFS), and overall survival (OS). Additionally, the addition of PD-L1 ICI to neoadjuvant chemotherapy has demonstrated the potential to achieve complete pathological response (pCR); and pCR has been associated with better long-term outcomes for locally advanced, resectable NSCLC. Regimens, including ICIs and TKIs, are now the standard of care for treatment in some early and resectable locally advanced stages of non-small cell lung cancer (NSCLC). Along with these advances and multiple therapeutic combinations, there is a need to find biomarkers that can identify the persistence of the disease or its recurrence and guide decision-making regarding the continuation of treatment, its suspension, or change.

The analysis of VOCs in exhaled breath is a promising non-invasive method for detecting and managing lung cancer. Several studies have explored the existence of a specific VOC profile among lung cancer patients. However, aims at finding specific metabolites frequently led to incoherent results (19). Indeed, if it would be possible to identify a specific compound or compounds pertaining to LC in exhaled air it would be a major improvement for LC screening and management. However, cancer is such a diverse disease that it is highly unlikely that a single biomarker or a small set of defined biomarkers will ever detect all cancers of a particular organ with high specificity and sensitivity. Instead, our previous work provided a different approach while showing a clear difference between the VOC profiles of lung cancer patients and healthy individuals without the qualitative information from specific compounds(26). The VOC profile reflects the cells’ metabolic activity, and it is expected that changes resulting from the presence or absence of the tumor will be displayed in the VOC profile.

By building on previous experience, this study aims to elucidate the alterations in the volatile organic compound (VOC) profile throughout the disease process of resectable, locally advanced NSCLC. Before initiating any therapeutic interventions, a comprehensive baseline VOC profile will be established for all enrolled patients. We hypothesize modifications in the VOC profile after well-succeeded surgical treatment and aim to establish the VOC profiles pattern along the follow-up time.

An interim analysis will be conducted during the follow-up period after an adequate number of patients have been enrolled in each study group, thereby facilitating data analysis execution. Numerous inquiries will be addressed throughout the study, including: In what ways do various treatments influence the VOC profile? Can it serve as a predictive tool for the recurrence of the disease in the absence of macroscopic evidence? What is the recurrence profile, and will it mirror the baseline established at the time of diagnosis? Will this change be sufficiently clear to enable a high degree of certainty regarding the recurrence or persistence of the disease? Ultimately, can this tool be used for screening and monitoring of lung cancer patients? For those patients included in Group S who are submitted exclusively to surgery, it is expected that tumor retrieval will significantly reduce the burden of disease and alter the VOC profile (VOC S_BL_ versus VOC _P_S). Will this profile be similar to that of healthy individuals (i.e., without any known previous neoplasic disease)? We will address this matter using the profile of healthy individuals established in our previous study (21). Over the two years of follow-up, will the profile remain unchanged if there is no evidence of recurrence?

For patients included in Group _N_S_A_ who are subjected to perioperative treatments, including ICIs, the pathologic response, especially for those patients with pCR, is expected to observe a change in the VOC profile. Therefore, VOC N_BL_ should be significantly different from VOC S_BL_. What will happen after surgery (VOC _P_S)? Will another change likely occur? If there is a pCR, will the VOC S_BL_ for Group _N_S_A_ be like the VOC _P_S for those patients in Group S? Does the pCR resulting from using ChT plus ICI have the same impact on the VOC profile as tumor retrieval surgery?

For patients experiencing an upstaging during surgery who will require adjuvant treatments (Group S_A_), the volatile organic compound (VOC) profile post-surgery (VOC _P_S) is anticipated to differ significantly from that of patients who do not experience such an upstaging. Specifically, in cases of upstaging, especially among those with lymph node involvement (N1 or N2), it is hypothesized that there exists an elevated risk of microscopic disease. Based on this hypothesis, it is expected that there will be discernible differences in the VOC profiles post-surgery (VOC _P_S) between the two groups (Group S versus Group S_A_).

Stratifying patients into different groups will allow establishing standard VOC profiles throughout the follow-up period. Variations to those patterns concomitantly confirmed with an imaging documented disease relapse would allow us to suggest that this method can effectively be useful in detecting LC recurrence *per se*.

The ultimate goal of this research is to validate the breath analysis as a reliable, rapid, non-invasive and more cost-effective approach for managing lung cancer in patients with an established diagnosis. The VOC profiling has the potential to be framed into a therapeutic and monitoring decision algorithm and at least complement imaging exams or even replace as a primary tool for disease monitoring in the future.

A new study will follow based on the same protocol but with a larger population and expanded to other clinical centers to validate the findings from the present study.

## Data Availability

This manuscript is a protocol for an ongoing study. No datasets were generated or analysed during the current study at this point. All relevant data from this study will be made available upon study completion.

